# Asymmetrical 5’RACE RNA-based immunoglobulins repertoire sequencing is an accurate and specific tool for diagnosis of POEMS syndrome

**DOI:** 10.1101/2024.11.15.24316857

**Authors:** Murielle Roussel, Virginie Pascal, Arnaud Jaccard, Frank Bridoux, Sebastien Bender, Christophe Sirac

## Abstract

POEMS syndrome is a rare form of plasma cell dyscrasia with multiple clinical features including polyneuropathy and sclerotic bone lesions. As various signs and symptoms can be observed, diagnosis is often delayed and we need an accurate diagnosis tool for these patients. Vascular endothelial growth factor (VEGF) is known to play a major role in the pathology and is frequently increased in sera of the patients but is not specific. This condition is also associated with the presence of a monoclonal immunoglobulin light chain composed of highly restricted variable domains (IGLV1-40, IGLV1-44, IGLV1-36 and IGJV3*02 genes). Using RNA-based high-throughput sequencing (5’ RACE RepSeq), we previously showed mutated consensus stretches in the CDR1/FR2 regions of the variable domains of the light chains: (P/A)VNWYQ and (D/G)VNWYQ. We have now added 70 light chain sequences to our cohort and confirmed the variable domain restriction, the consensus stretches and found a new YSVNWYQ pattern on IGLV1-44 light chains. When correlated with our clinical database of POEMS patients, these mutation patterns had an accuracy of 89% and a specificity of 100%. 5’RACE RepSeq is an important tool to ensure the diagnosis of patients suspected of having POEMS syndrome, even with a small secretory clone. Samples from an involved organ are mandatory in the case of localized disease.

## INTRODUCTION

POEMS is a rare paraneoplastic syndrome associated with an underlying plasma cell dyscrasia. It is characterized by Peripheral neuropathy, Organomegaly, Endocrinopathy, Monoclonal component, Skin changes, papillary edema, extravascular volume overload, sclerotic bone lesions, pulmonary dysfunction (restrictive syndrome or pulmonary hypertension), and thrombocytosis or erythrocytosis^1,2^. A. Dispenzieri and colleagues defined, more than 15 years ago, diagnosis criteria^3,4^ that are widely accepted but, as various signs and symptoms can be related to that condition, diagnosis is often delayed with impaired neurological functions and there is still an unmet need for an accurate diagnosis tool. Although the physiopathology remains elusive, the presence of a monoclonal lambda light chain deriving mainly from 2 variable germline genes, IGLV1-40 or IGLV1-44^5^ and the overproduction of cytokines, particularly vascular endothelial growth factor (VEGF)^6–9^, likely play a major role in that disorder. We previously reported recurrent mutations in 3 variable (V_L_) segments of lambda light chains (IGLV1-40,1-44 and 1-36), all associated with an IGLJ3*02 junction (J) gene, confirming a highly restrictive IGLV-J germline gene usage in POEMS syndrome^5,10–12^.

High-throughput repertoire sequencing on RNA with 5’Rapid Amplification of cDNA Ends-based (5’RACE RepSeq) is a sensitive technique that was developed to estimate the dominant immune repertoire diversity^13 14^. We applied that technique to sequence the immunoglobulins repertoire providing full-length V(D)J regions sequences for the analysis of immunoglobulin variable gene frequencies, mutational patterns and diversity due to somatic hypermutation. To evaluate the accuracy of the technique as a specific diagnosis test in POEMS patients, we prospectively analyzed since 2018 mRNA encoding immunoglobulins in 131 samples (bone marrow aspirate or biopsies, lymph nodes biopsies, ascites aspirates) from 114 patients with suspected POEMS syndrome. We were able to review complete clinical, and biological data in 91% of patients resulting in a truthful database of 105 patients with suspected POEMS syndrome. We correlated our results of immunoglobulins repertoire sequencing to the clinical data to evaluate the accuracy, sensitivity and specificity of this diagnosis tool.

## METHODS

RNA sample preparation and High-throughput sequencing on Illumina MiSeq sequencer.

Total RNAs were extracted from fresh BM/ascites aspirates or bone/lymph node biopsies, after collection in RNA preservation tubes containing cell lysis and RNA stabilization reagents (Tempus RNA Tubes (ThermoFisher) for aspirates and RNA later (Invitrogen) for biopsies). Complementary DNAs were obtained by asymmetrical 5’RACE/reverse transcription with five antisense specific primers for the constant regions of IGH or IGK/L (C*μ*, C*γ*, C*α*, C*κ*, or C*λ*), a 5’ end CCCC tag, and a universal 3’ forward primer GGGG coupled to unique molecular identifiers (UMIs) in order to allow to trace reads back to their original cDNA molecules thus correcting for sequencing errors. Initial amplification relies on semi-nested PCR with a nested reverse primer specific to the first constant domain CH1 for heavy and light chains, followed by a series of PCRs for paired-end sequencing using the Illumina® V3 and V2 MiSeq kit, with asymmetric reading to obtain sequences up to 750 nucleotides (entire variable domain and part of the constant region) and on average 325000 reads per sample. Repertoire analysis was performed using the VIDJIL tool (http://vidjil.org/)^15^ and international ImMunoGenetics (IMGT)/HIGHV-QUEST tool (http://imgt.org/), resulting in consensus sequences for each clonotype^16^. Initial biological data are available at European Nucleotide Archive (ENA) database under accession number PRJEB35157. All data produced in the present study are available upon reasonable request to the authors. Patients signed the informed consent for the RNA sequencing of their immunoglobulins’ repertoire. The database is declared at the French Commission Nationale de l’Informatique et des Libertés (CNIL DR211392) and the Comité Consultatif sur le Traitement de l’Information en Matière de Recherche dans le Domaine de la Santé (CCTIRS N°1158) gave approvals for additional data processing. This study has received approval for retention and treatment of biological samples from the Comité de Protection des Personnes (CPP DC-2008-111). Research was conducted in accordance with the Declaration of Helsinki.

## RESULTS AND DISCUSSION

5’RACE RepSeq detected “dominant” clonotypes in the repertoire of immunoglobulins of 86/105 (82%) patients. The distribution of main IGK/LV-J variable domains showed, as expected, an IGLV-J predominance as all but six patients referred for a suspected POEMS syndrome had a monoclonal lambda light chain. Overall, 80/86 (93%) of suspected POEMS patients had a lambda clone with either IGLV1-44 (n=35, 41%), IGLV1-40 (n=16, 18.5%), or IGLV1-36 (n=4, 4.5%) variable genes for 55 of them and a germline IGLJ3*02 junction gene in 57 (66%) pts. Fourteen patients had 2 dominant clonotypes. We confirmed the presence of the recurrent consensus stretches *(P/A/D/G)VNWYQ* in the CDR1, FR2 regions according to the IMGT unique numbering^16^ in 45 patients including the following mutations: IGLV1-44 T38>P n=18, IGLV1-44 T38>A n=9, IGLV1-40 H40>N n=12, and IGLV1-44 T38>G n=2, IGLV1-36 A38>G n=3. The addition of new clinically documented patients (n=70) allowed us to describe a new motif in 3 patients with IGLV1-44 sequences comprising N37>Y and T38>S substitutions (*YSVNWYQ*), mutations that are highly dissimilar to the germline NT motif according to IMGT physicochemical classes of AA. Seven patients had no mutation (n=3) or non-classical AA substitutions/mutations (T38>Y, T38>S, and H40>Q), only 2 of them were IGLJ3*02. Alignments of AA with colored substitutions/mutations of the IGLV-J sequences are shown in ***Fig. 1***.

**Fig. 1.**
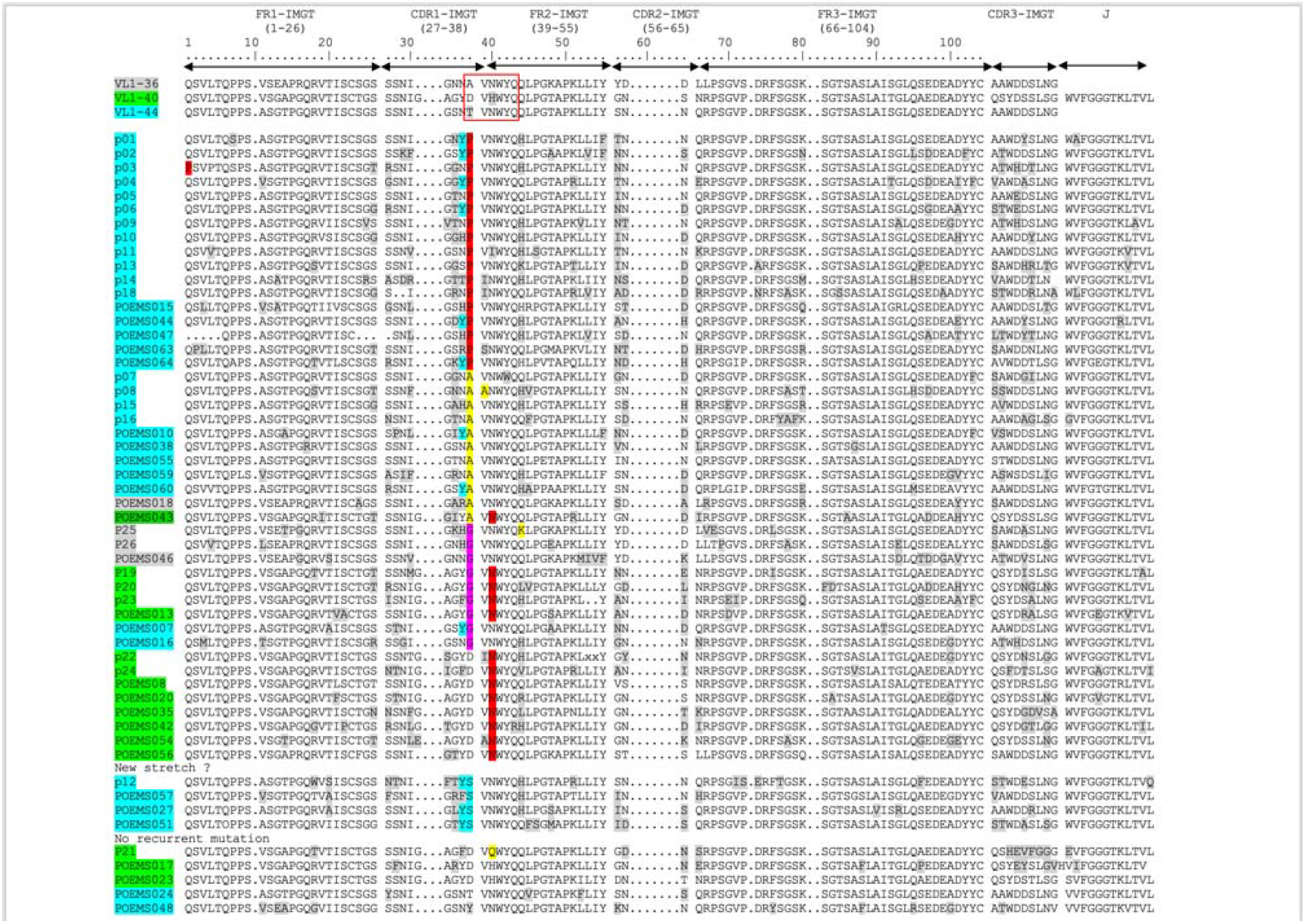
AA Alignments of POEMS sequences. Deduced AA sequences of the monoclonal IGLV-J domains in patients with POEMS syndrome compared with germline sequences according to IMGT numbering.

**Fig. 2.**
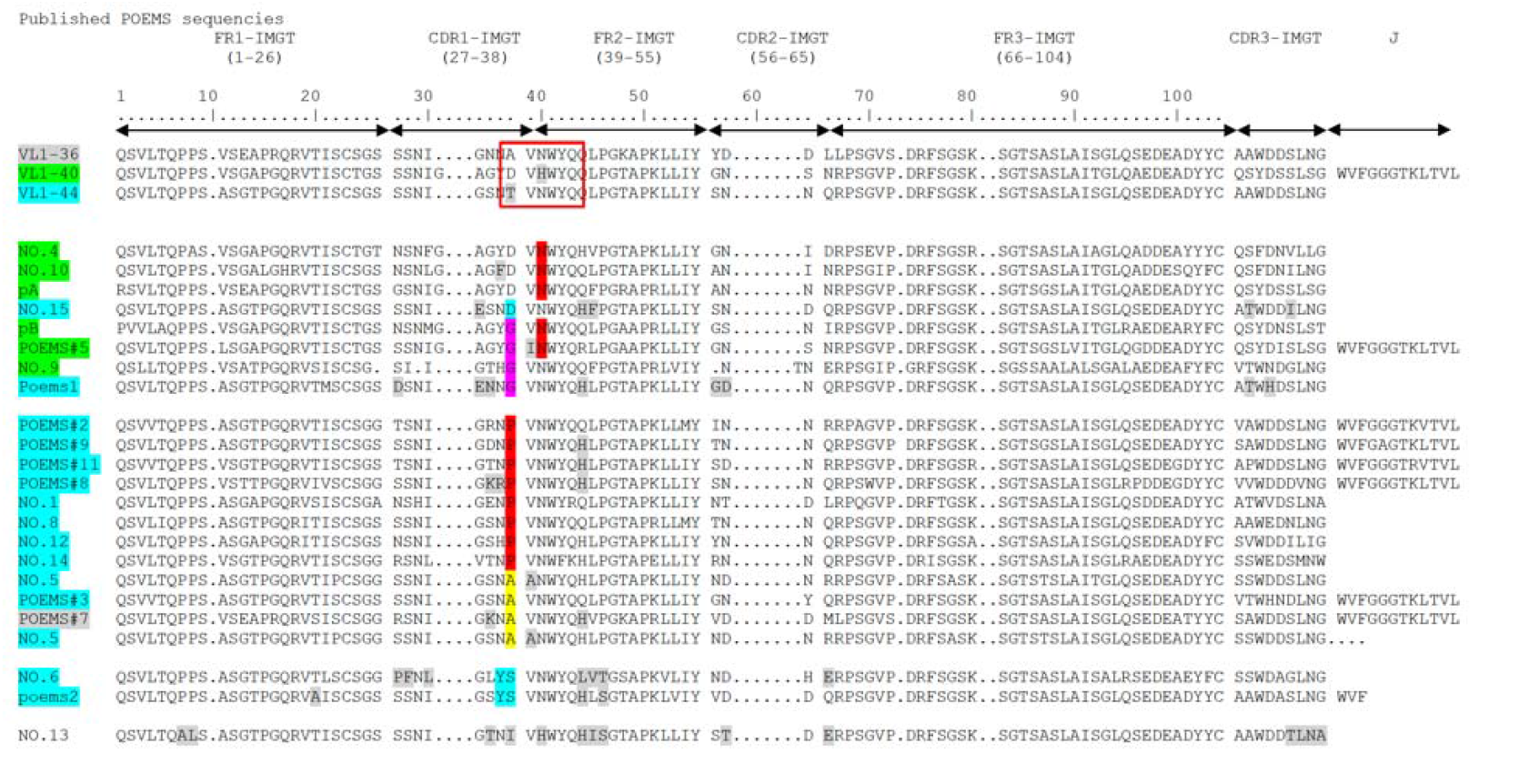
Other Published POEMS sequences (from AL-Base^17^) Deduced AA published sequences of the monoclonal IGLV-J domains in patients with POEMS syndrome.

We subsequently used the same IGLV1-J3*02 sequences of our own immunoglobulins related diseases (MGCS/MGRS/AL/MM) database (n=92/396) and the online AL-Base database^17^ to confirm the specificity of these patterns. All referenced POEMS patients (excluding our previously published sequencies) had the specific stretches *(P/A)VNWYQ, (D/G)VNWYQ* in the IGLV1-40, 1-44 or 1-36 variable genes. Two published POEMS patients (NO.6^5^, POEMS2^10^) had an YS motif. Screening for these mutational patterns was negative within our non-POEMS immunoglobulins sequences and we only found 1 patient in the whole AL-Base over the 276 listed IGLV1 monoclonal gammopathies sequences, with either the *(P/A)VNWYQ, (D/G)VNWYQ or YSVNWYQ* stretches and the IGLJ3*02 gene (AL amyloidosis patient with no available clinical data, seq. GET: IGLV1-40,J3*02). To determine the accuracy, sensibility and specificity of these recurrent consensus stretches detected by 5’RACE RepSeq, we correlated the IGLV-J mutational patterns to the patients’ clinical data. According to diagnosis criteria and regardless of the IGVL-J patterns, the diagnosis of POEMS syndrome was certain for 67 patients, probable for 6, and possible for 3 of them. Overall, 29 patients did not actually meet the criteria for POEMS syndrome. Clinical and biological characteristics of the patients are shown in ***Table 1***.

**Table 1.**
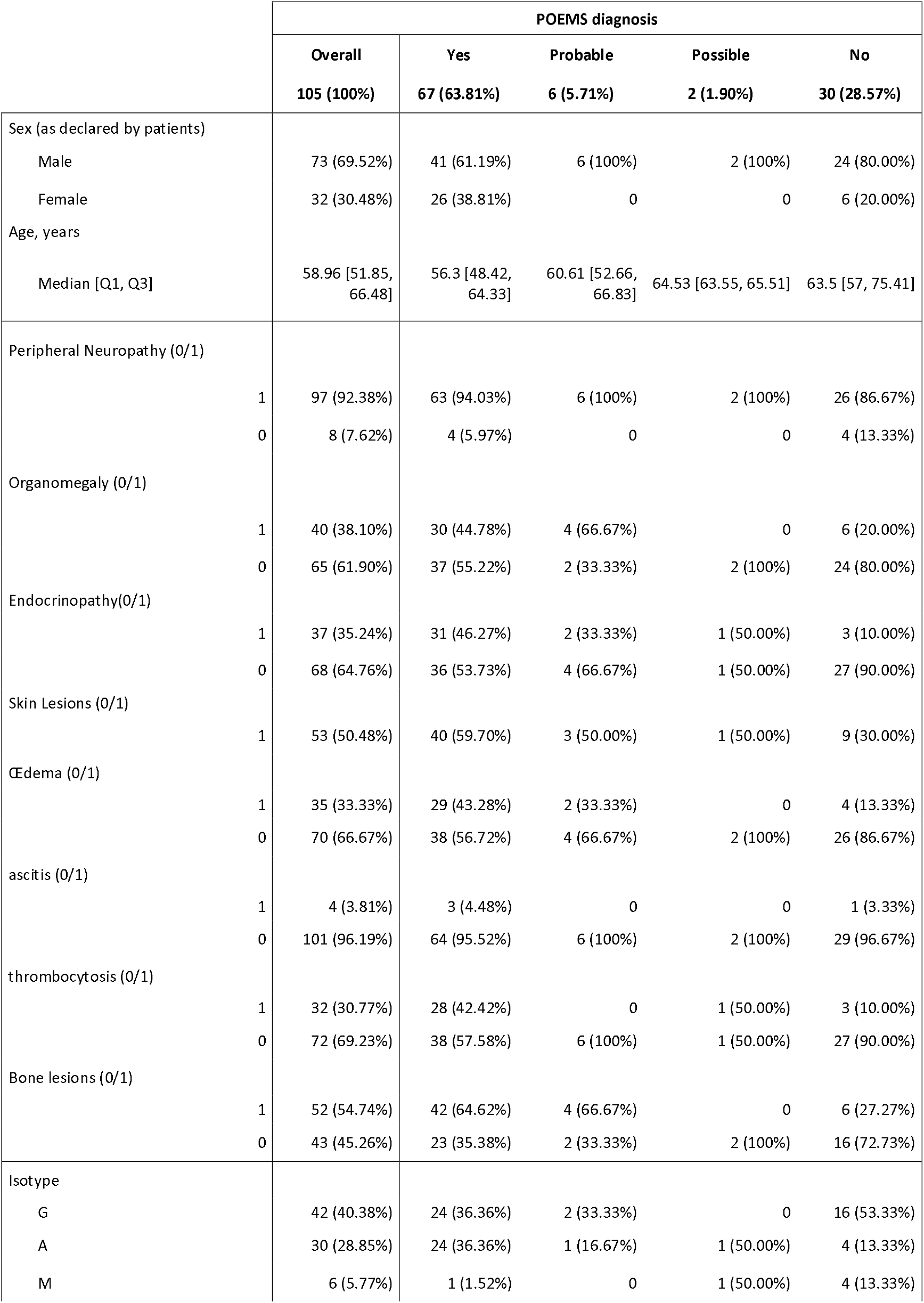
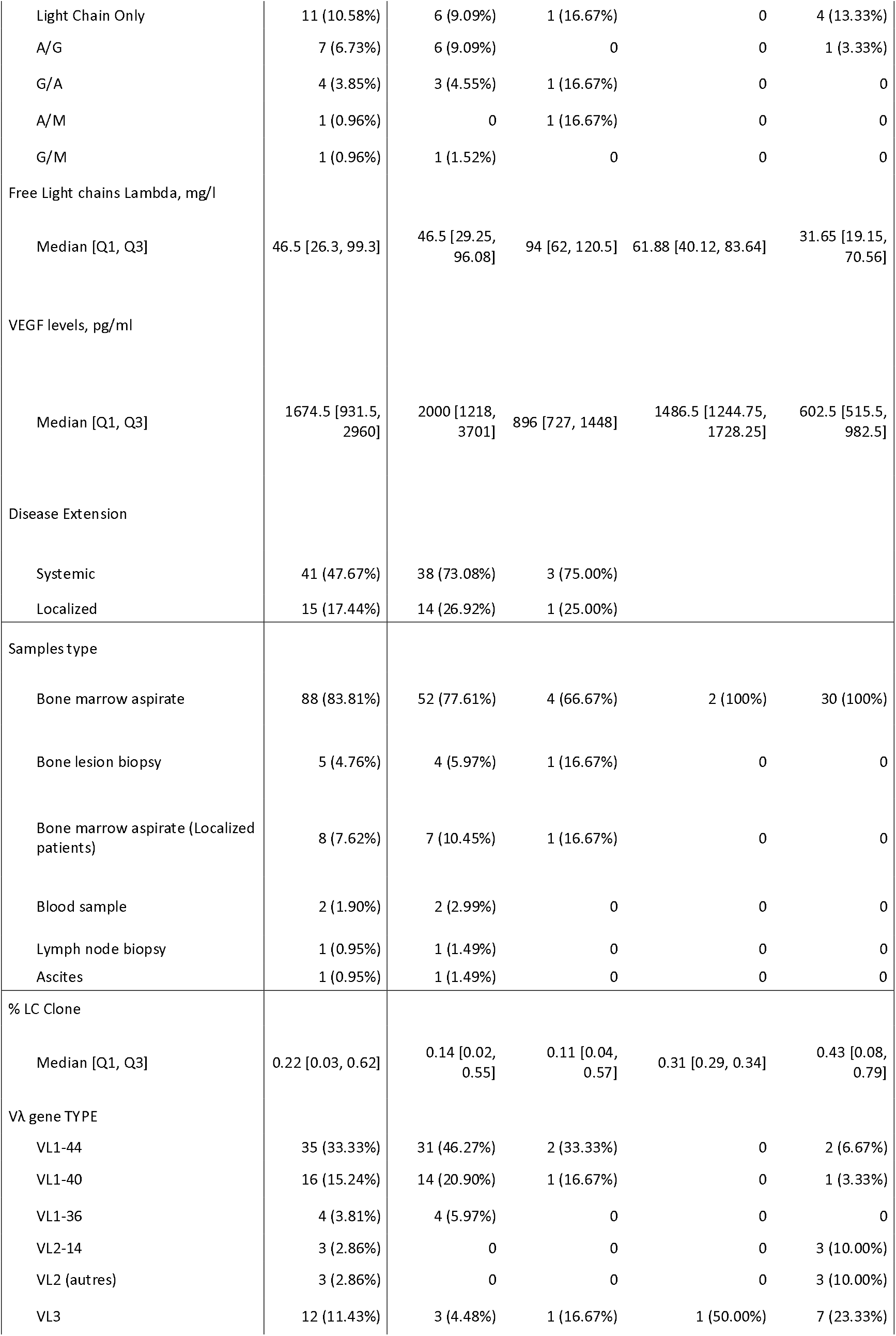
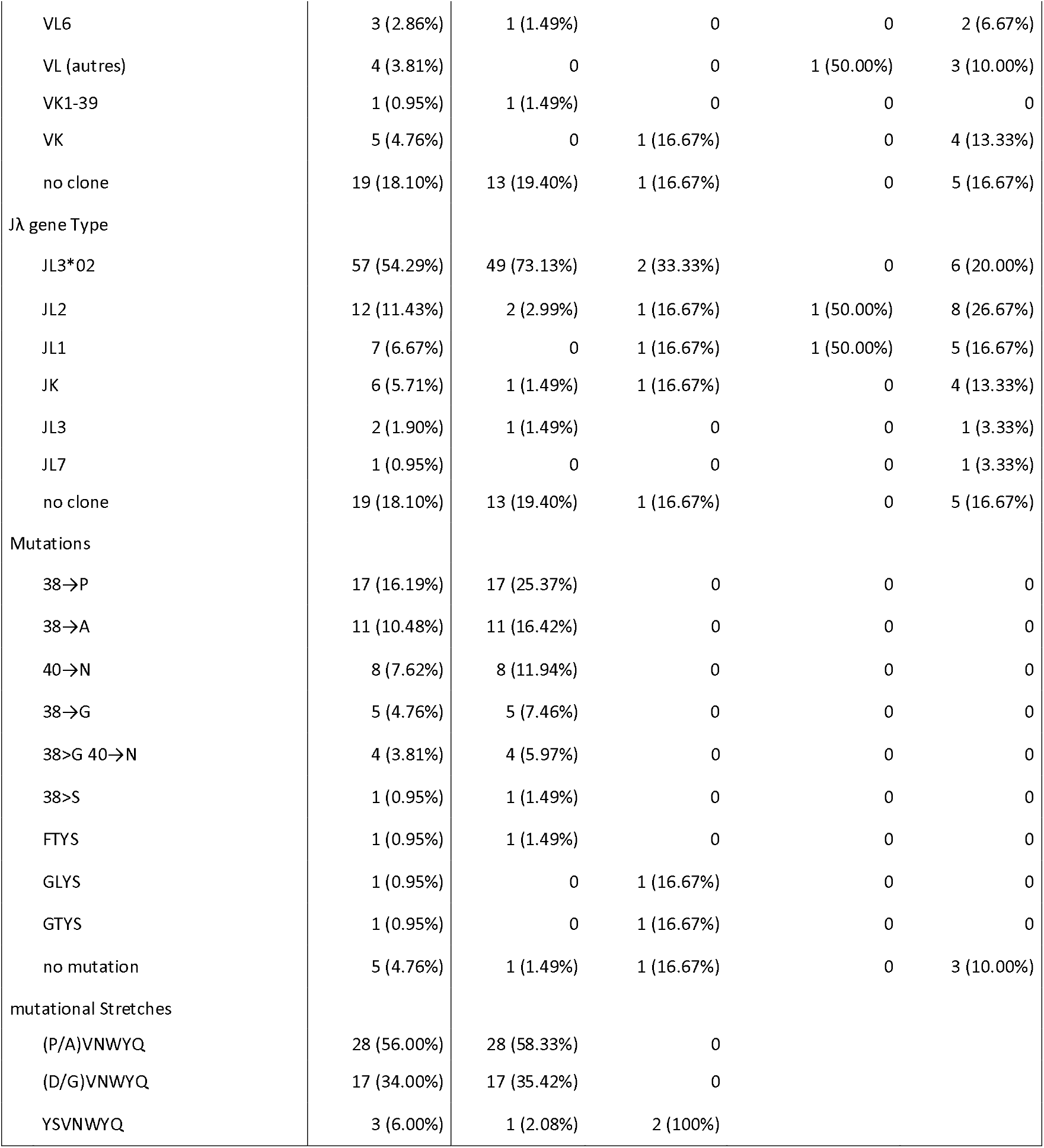
Clinical and biological characteristics of patients with suspected POEMS syndrome.

|  |  | POEMS diagnosis |  |  |  |  |
| --- | --- | --- | --- | --- | --- | --- |
|  |  | Overall<br>105 (100%) | Yes<br>67 (63.81%) | Probable<br>6 (5.71%) | Possible<br>2 (1.90%) | No<br>30 (28.57%) |
| Sex (as declared by patients) |  |  |  |  |  |  |
| Male |  | 73 (69.52%) | 41 (61.19%) | 6 (100%) | 2 (100%) | 24 (80.00%) |
| Female |  | 32 (30.48%) | 26 (38.81%) | 0 | 0 | 6 (20.00%) |
| Age, years |  |  |  |  |  |  |
| Median [Q1, Q3] |  | 58.96 [51.85, 66.48] | 56.3 [48.42, 64.33] | 60.61 [52.66, 66.83] | 64.53 [63.55, 65.51] | 63.5 [57, 75.41] |
| Peripheral Neuropathy (0/1) |  |  |  |  |  |  |
|  | 1 | 97 (92.38%) | 63 (94.03%) | 6 (100%) | 2 (100%) | 26 (86.67%) |
|  | 0 | 8 (7.62%) | 4 (5.97%) | 0 | 0 | 4 (13.33%) |
| Organomegaly (0/1) |  |  |  |  |  |  |
|  | 1 | 40 (38.10%) | 30 (44.78%) | 4 (66.67%) | 0 | 6 (20.00%) |
|  | 0 | 65 (61.90%) | 37 (55.22%) | 2 (33.33%) | 2 (100%) | 24 (80.00%) |
| Endocrinopathy(0/1) |  |  |  |  |  |  |
|  | 1 | 37 (35.24%) | 31 (46.27%) | 2 (33.33%) | 1 (50.00%) | 3 (10.00%) |
|  | 0 | 68 (64.76%) | 36 (53.73%) | 4 (66.67%) | 1 (50.00%) | 27 (90.00%) |
| Skin Lesions (0/1) |  |  |  |  |  |  |
|  | 1 | 53 (50.48%) | 40 (59.70%) | 3 (50.00%) | 1 (50.00%) | 9 (30.00%) |
| Œdema (0/1) |  |  |  |  |  |  |
|  | 1 | 35 (33.33%) | 29 (43.28%) | 2 (33.33%) | 0 | 4 (13.33%) |
|  | 0 | 70 (66.67%) | 38 (56.72%) | 4 (66.67%) | 2 (100%) | 26 (86.67%) |
| ascitis (0/1) |  |  |  |  |  |  |
|  | 1 | 4 (3.81%) | 3 (4.48%) | 0 | 0 | 1 (3.33%) |
|  | 0 | 101 (96.19%) | 64 (95.52%) | 6 (100%) | 2 (100%) | 29 (96.67%) |
| thrombocytosis (0/1) |  |  |  |  |  |  |
|  | 1 | 32 (30.77%) | 28 (42.42%) | 0 | 1 (50.00%) | 3 (10.00%) |
|  | 0 | 72 (69.23%) | 38 (57.58%) | 6 (100%) | 1 (50.00%) | 27 (90.00%) |
| Bone lesions (0/1) |  |  |  |  |  |  |
|  | 1 | 52 (54.74%) | 42 (64.62%) | 4 (66.67%) | 0 | 6 (27.27%) |
|  | 0 | 43 (45.26%) | 23 (35.38%) | 2 (33.33%) | 2 (100%) | 16 (72.73%) |
| Isotype |  |  |  |  |  |  |
| G |  | 42 (40.38%) | 24 (36.36%) | 2 (33.33%) | 0 | 16 (53.33%) |
| A |  | 30 (28.85%) | 24 (36.36%) | 1 (16.67%) | 1 (50.00%) | 4 (13.33%) |
| M |  | 6 (5.77%) | 1 (1.52%) | 0 | 1 (50.00%) | 4 (13.33%) |

|  |  |  |  |  |  |
| --- | --- | --- | --- | --- | --- |
| Light Chain Only | 11 (10.58%) | 6 (9.09%) | 1 (16.67%) | 0 | 4 (13.33%) |
| A/G | 7 (6.73%) | 6 (9.09%) | 0 | 0 | 1 (3.33%) |
| G/A | 4 (3.85%) | 3 (4.55%) | 1 (16.67%) | 0 | 0 |
| A/M | 1 (0.96%) | 0 | 1 (16.67%) | 0 | 0 |
| G/M | 1 (0.96%) | 1 (1.52%) | 0 | 0 | 0 |
| Free Light chains Lambda, mg/l |  |  |  |  |  |
| Median [Q1, Q3] | 46.5 [26.3, 99.3] | 46.5 [29.25, 96.08] | 94 [62, 120.5] | 61.88 [40.12, 83.64] | 31.65 [19.15, 70.56] |
| VEGF levels, pg/ml |  |  |  |  |  |
| Median [Q1, Q3] | 1674.5 [931.5, 2960] | 2000 [1218, 3701] | 896 [727, 1448] | 1486.5 [1244.75, 1728.25] | 602.5 [515.5, 982.5] |
| Disease Extension |  |  |  |  |  |
| Systemic | 41 (47.67%) | 38 (73.08%) | 3 (75.00%) |  |  |
| Localized | 15 (17.44%) | 14 (26.92%) | 1 (25.00%) |  |  |
| Samples type |  |  |  |  |  |
| Bone marrow aspirate | 88 (83.81%) | 52 (77.61%) | 4 (66.67%) | 2 (100%) | 30 (100%) |
| Bone lesion biopsy | 5 (4.76%) | 4 (5.97%) | 1 (16.67%) | 0 | 0 |
| Bone marrow aspirate (Localized patients) | 8 (7.62%) | 7 (10.45%) | 1 (16.67%) | 0 | 0 |
| Blood sample | 2 (1.90%) | 2 (2.99%) | 0 | 0 | 0 |
| Lymph node biopsy | 1 (0.95%) | 1 (1.49%) | 0 | 0 | 0 |
| Ascites | 1 (0.95%) | 1 (1.49%) | 0 | 0 | 0 |
| % LC Clone |  |  |  |  |  |
| Median [Q1, Q3] | 0.22 [0.03, 0.62] | 0.14 [0.02, 0.55] | 0.11 [0.04, 0.57] | 0.31 [0.29, 0.34] | 0.43 [0.08, 0.79] |
| Vλ gene TYPE |  |  |  |  |  |
| VL1-44 | 35 (33.33%) | 31 (46.27%) | 2 (33.33%) | 0 | 2 (6.67%) |
| VL1-40 | 16 (15.24%) | 14 (20.90%) | 1 (16.67%) | 0 | 1 (3.33%) |
| VL1-36 | 4 (3.81%) | 4 (5.97%) | 0 | 0 | 0 |
| VL2-14 | 3 (2.86%) | 0 | 0 | 0 | 3 (10.00%) |
| VL2 (autres) | 3 (2.86%) | 0 | 0 | 0 | 3 (10.00%) |
| VL3 | 12 (11.43%) | 3 (4.48%) | 1 (16.67%) | 1 (50.00%) | 7 (23.33%) |
| VL6 | 3 (2.86%) | 1 (1.49%) | 0 | 0 | 2 (6.67%) |
| VL (autres) | 4 (3.81%) | 0 | 0 | 1 (50.00%) | 3 (10.00%) |
| VK1-39 | 1 (0.95%) | 1 (1.49%) | 0 | 0 | 0 |
| VK | 5 (4.76%) | 0 | 1 (16.67%) | 0 | 4 (13.33%) |
| no clone | 19 (18.10%) | 13 (19.40%) | 1 (16.67%) | 0 | 5 (16.67%) |
| Jλ gene Type |  |  |  |  |  |
| JL3*02 | 57 (54.29%) | 49 (73.13%) | 2 (33.33%) | 0 | 6 (20.00%) |
| JL2 | 12 (11.43%) | 2 (2.99%) | 1 (16.67%) | 1 (50.00%) | 8 (26.67%) |
| JL1 | 7 (6.67%) | 0 | 1 (16.67%) | 1 (50.00%) | 5 (16.67%) |
| JK | 6 (5.71%) | 1 (1.49%) | 1 (16.67%) | 0 | 4 (13.33%) |
| JL3 | 2 (1.90%) | 1 (1.49%) | 0 | 0 | 1 (3.33%) |
| JL7 | 1 (0.95%) | 0 | 0 | 0 | 1 (3.33%) |
| no clone | 19 (18.10%) | 13 (19.40%) | 1 (16.67%) | 0 | 5 (16.67%) |
| Mutations |  |  |  |  |  |
| 38→P | 17 (16.19%) | 17 (25.37%) | 0 | 0 | 0 |
| 38→A | 11 (10.48%) | 11 (16.42%) | 0 | 0 | 0 |
| 40→N | 8 (7.62%) | 8 (11.94%) | 0 | 0 | 0 |
| 38→G | 5 (4.76%) | 5 (7.46%) | 0 | 0 | 0 |
| 38>G 40→N | 4 (3.81%) | 4 (5.97%) | 0 | 0 | 0 |
| 38>S | 1 (0.95%) | 1 (1.49%) | 0 | 0 | 0 |
| FTYS | 1 (0.95%) | 1 (1.49%) | 0 | 0 | 0 |
| GLYS | 1 (0.95%) | 0 | 1 (16.67%) | 0 | 0 |
| GTYS | 1 (0.95%) | 0 | 1 (16.67%) | 0 | 0 |
| no mutation | 5 (4.76%) | 1 (1.49%) | 1 (16.67%) | 0 | 3 (10.00%) |
| mutational Stretches |  |  |  |  |  |
| (P/A)VNWYQ | 28 (56.00%) | 28 (58.33%) | 0 |  |  |
| (D/G)VNWYQ | 17 (34.00%) | 17 (35.42%) | 0 |  |  |
| YSVNWYQ | 3 (6.00%) | 1 (2.08%) | 2 (100%) |  |  |

Regarding the 19 patients with no medullary dominant clonotype detected, 13 patients had an overt POEMS and one patient has a probable POEMS syndrome. Of importance, 8 of them had a localized disease with no patent bone marrow involvement that could explain the negativity of the bone marrow samples and 2 further patients’ samples were actually blood aspirates. Within the 6 patients with only a dominant IGKV clone detected, 2 had a diagnosis of POEMS certain or probable. Therefore, 80 5’RACE RepSeq/clinical data couples were analyzed. We confirmed that 5’RACE RepSeq is an accurate diagnostic tool for POEMS syndrome with a specificity of 100%, sensibility of 79%, and accuracy of 85% ***(Table 2)***: the specific consensus stretches in the IGLV-J sequences were detected only in patients with clinically confirmed POEMS syndrome. If we add the 3 YS patients (not previously considered as specific), all were confirmed or probable POEMS, raising accuracy to 89%, and sensitivity to 84% while maintaining specificity at 100%. We were not able to distinguish clinical phenotypes according to the mutational stretches.

**Table 2.** Contingency tables for 5’RACE RepSeq according to clinical data.

|  |  | POEMS |  |  |  |
| --- | --- | --- | --- | --- | --- |
|  |  | YES ( 1 and 1_2) | NO (0 and 2) |  |  |
| Typical Stretches | YES | 45 | 0 | 1 | PPV |
|  | NO | 12 | 23 | 0,65714286 | NPV |
|  |  | 0,789473684 | 1 | 0,85 | Accuracy |
|  |  | Sensibility | Specificity |  |  |

B: NGS with (P/A)VNWYQ, (D/G)VNWYQ, and YSVNWYQ
|  |  | POEMS |  |  |  |
| --- | --- | --- | --- | --- | --- |
|  |  | YES ( 1 and 1_2) | NO (0 and 2) |  |  |
| Typical Stretches | YES | 48 | 0 | 1 | PPV |
|  | NO | 9 | 23 | 0,71875 | NPV |
|  |  | 0,842105263 | 1 | 0,8875 | Accuracy |
|  |  | Sensibility | Specificity |  |  |

PPV: Predictive Positive Value; NPV: Negative Predictive Value

Our results confirmed that asymmetrical 5’RACE RepSeq is an 89% accurate diagnosis tool for POEMS syndrome with 100% specificity and 84% sensitivity. It can detect “pathological” clones with typical stretches *(P/A)VNWYQ, (D/G)VNWYQ*, and *YSVNWYQ* in the IGLV1-40, 1-44 or 1-36 variable genes, but, in case of localized disease with no “systemic” bone marrow involvement, samples of an involved organ are mandatory (lymph node, sclerotic bone lesion, ascites…). We will further study whether the clonotypic identification of the IGLV-J sequence could allow us to follow POEMS patients in blood during therapy and/or remission through clonotypic peptides Mass Spectrometry.

## Data Availability

All data produced in the present study are available upon reasonable request to the authors

## Acknowledgements

The authors gratefully acknowledge individual physicians and technical teams at all centers and especially all members of the French Network for immunoglobulins related disorders chaired by Pr Arnaud JACCARD and Frank BRIDOUX. This work was funded by Foundation Française pour la Recherche sur le Myélome et les Gammapathies Monoclonales.

## Authorship Contributions

SB and VP performed the 5’RACE RepSeq; MR, VP, AJ, FB and SB collected and analyzed the data, MR, SB, and CS wrote the paper.

## REFERENCES

1. Miralles GD, O’Fallon JR, Talley NJ. Plasma-cell dyscrasia with polyneuropathy. The spectrum of POEMS syndrome. N Engl J Med 1992;327(27):1919–23.

2. Bardwick PA, Zvaifler NJ, Gill GN, Newman D, Greenway GD, Resnick DL. Plasma cell dyscrasia with polyneuropathy, organomegaly, endocrinopathy, M protein, and skin changes: the POEMS syndrome. Report on two cases and a review of the literature. Medicine (Baltimore) 1980;59(4):311–22.

3. Dispenzieri A. POEMS syndrome: Update on diagnosis, risk-stratification, and management. Am J Hematol 2023;98(12):1934–50.

4. Dispenzieri A, Kyle RA, Lacy MQ, et al. POEMS syndrome: definitions and long-term outcome. Blood 2003;101(7):2496–506.

5. Abe D, Nakaseko C, Takeuchi M, et al. Restrictive usage of monoclonal immunoglobulin lambda light chain germline in POEMS syndrome. Blood 2008;112(3):836–9.

6. D’Souza A, Hayman SR, Buadi F, et al. The utility of plasma vascular endothelial growth factor levels in the diagnosis and follow-up of patients with POEMS syndrome. Blood 2011;118(17):4663–5.

7. Watanabe O, Arimura K, Kitajima I, Osame M, Maruyama I. Greatly raised vascular endothelial growth factor (VEGF) in POEMS syndrome. Lancet 1996;347(9002):702.

8. Watanabe O, Maruyama I, Arimura K, et al. Overproduction of vascular endothelial growth factor/vascular permeability factor is causative in Crow-Fukase (POEMS) syndrome. Muscle Nerve 1998;21(11):1390–7.

9. Gherardi RK, Bélec L, Soubrier M, et al. Overproduction of proinflammatory cytokines imbalanced by their antagonists in POEMS syndrome. Blood 1996;87(4):1458–65.

10. Kawajiri-Manako C, Mimura N, Fukuyo M, et al. Clonal immunoglobulin λ light-chain gene rearrangements detected by next generation sequencing in POEMS syndrome. Am J Hematol 2018;93(9):1161–8.

11. Bender S, Javaugue V, Saintamand A, et al. Immunoglobulin variable domain high-throughput sequencing reveals specific novel mutational patterns in POEMS syndrome. Blood 2020;135(20):1750–8.

12. Isshiki Y, Oshima M, Mimura N, et al. Unraveling unique features of plasma cell clones in POEMS syndrome with single-cell analysis. JCI Insight 2022;7(20):e151482.

13. van Dongen JJM, Langerak AW, Brüggemann M, et al. Design and standardization of PCR primers and protocols for detection of clonal immunoglobulin and T-cell receptor gene recombinations in suspect lymphoproliferations: report of the BIOMED-2 Concerted Action BMH4-CT98-3936. Leukemia 2003;17(12):2257–317.

14. Calis JJA, Rosenberg BR. Characterizing immune repertoires by high throughput sequencing: strategies and applications. Trends Immunol 2014;35(12):581–90.

15. Vidjil: A Web Platform for Analysis of High-Throughput Repertoire Sequencing | PLOS ONE https://journals-plosorg.proxy.insermbiblio.inist.fr/plosone/article?id=10.1371/journal.pone.0166126

16. Alamyar E, Duroux P, Lefranc M-P, Giudicelli V. IMGT® Tools for the Nucleotide Analysis of Immunoglobulin (IG) and T Cell Receptor (TR) V-(D)-J Repertoires, Polymorphisms, and IG Mutations: IMGT/V-QUEST and IMGT/HighV-QUEST for NGS. In: Christiansen FT, Tait BD, editors. Immunogenetics: Methods and Applications in Clinical Practice. Totowa, NJ: Humana Press; 2012. p. 569–604.Available from: 10.1007/978-1-61779-842-9_32

17. Bodi K, Prokaeva T, Spencer B, Eberhard M, Connors LH, Seldin DC. AL-Base: a visual platform analysis tool for the study of amyloidogenic immunoglobulin light chain sequences. Amyloid 2009;16(1):1–8.

